# Vaccine hesitancy for coronavirus SARS-CoV-2 in North India

**DOI:** 10.1101/2021.10.24.21265455

**Authors:** Utkarsh Srivastava, Avanish Kumar Tripathi, Jagjeet Kaur, Sabita Devi, Shipra Verma, Vanya Singh, Debashruti Das, Prajjval Pratap Singh, Pradeep Kumar, Vandana Rai, Rakesh Pandey, Gyaneshwer Chaubey

## Abstract

With the roll-out of world’s largest vaccine drive for Severe Acute Respiratory Syndrome coronavirus 2 (SARS-CoV-2) by Government of India on January 16 2021, India has targeted to vaccinate its entire population by the end of year 2021. Struggling with vaccine procurement and production earlier, India came up with these hurdles but the Indian population still did not seem to be mobilizing swiftly towards vaccination centers. With the initial hesitancy, as soon as the vaccination started to speedup, India was hit severely by the second wave. The severe second wave has slowed down the vaccination pace and also it was one of the major contributing factor of vaccine hesitancy. To understand the nature of vaccine hesitancy and factors underlying it, we conducted an extensive online and offline surveys in Varanasi and adjoining regions using structured questions. Majority of respondents though were students (0.633), respondents from other occupations such as government officials (0.10) were also included in the study. We observed several intriguing opinions on our eleven questions. It is interesting to note that the majority of the people (0.75) relied on fake news and did not take COVID-19 seriously. Most importantly, we noticed that a substantial proportion of respondents (relative frequency 0.151; mean age 24.8 years) reported that they are still not interested in vaccination. People who have neither been vaccinated nor have ever been infected may become the medium for spreading the virus and creating new variants. This could also lead to a resistant variant of the vaccine in the future. We expect that this extensive survey may help the government to upgrade their vaccination policies for COVID-19 in North India.

## INTRODUCTION

COVID-19 has impacted our life in multiple ways (*1, 2*). Since this disease is new, the information related to it is not very concrete. With the new researches accumulating everyday (*3*), the WHO as well as government guidelines have changed substantially. These changes have added mystification to the general populations (*4*–*6*). Thus, several local rumors against the vaccination drive have been developed in the population (*7, 8*). Since, the flow of information in India society is heavily dependent upon oral transmission i.e., world-of-mouth, many people are afraid to visit the vaccination centers (*9*).

India has begun the vaccination drive from 16^th^ January 2021. This was the time when only ∼200,000 cases were active and most Indians have overcome the trauma of the first wave (*10*). With the repeated encouragement from the government, India has achieved 22 million doses per day at the end of March 2021 (*11*). This number has increased exponentially during first week of April 2021, when government has decided to vaccinate everyone above 45 years of the age (*12*). However, this was also the time of the beginning of the second wave (*13, 14*). Due to the severe second wave the numbers of daily vaccine doses administered, which were more than 35 million a day till 13^th^ April 2021, have reduced to less than 15 million a day after a month (*11*). Moreover, leaders from several political parties have released public statements against the vaccination (*15*). The aforesaid appears to be major contributors to the reduced rate of vaccination after second wave (10). However, to the best of our knowledge any systematic effort to uncover the cause of vaccine hesitancy among Indians has not been made. Some empirical evidence is much needed to understand the nature and cause of the vaccine hesitancy in order to suggest potential psychosocial intervention to help the Indian policy makers and immunization staff to overcome this major hurdle in immunization against COVID-19.

To understand the nature and causes of vaccine hesitancy among Indians we conducted an extensive survey in Varanasi and adjoining regions (North India). To uncover the factors that are making Indian people hesitant towards taking the vaccine of SARS-CoV-2 and how socio-political aspects have affected the vaccination drive and shaped people’s views, we followed questionnaire based survey approach where we presented structured questions with predefined set of responses for each question.

## Methodology

### Participants

The study was conducted on relatively large incidental sample of participants (N =603 Males =337, Females= 266) in the age range of 18 to 40 years (mean age =26.9; SD=4.4). Only those respondents were included in the study who volunteered themselves and consented to take part in the study. Together with the online survey, we have also conducted an offline survey (telephonic interview). For the analysis, we have anonymized the participants. The study has been approved by the Ethical committees of Banaras Hindu University, Varanasi and VBS Purvanchal University, Jaunpur, India. Though attempt was made to recruit participants from different occupational background, majority of respondents were students (0.633) with relatively few government employees (0.10).

### Materials and procedure

We conducted a questionnaire based survey (supplementary text) consisting of 11 questions related to awareness about COVID-19 pandemic, its spread and vaccine. The survey was though largely conducted through online platform, the telephonic survey was also done to readily reach the people from the rural areas (who were not able to use online platform) and know their attitudes and perspectives regarding the whole COVID-19 scenario (which is equally important as that of urban people). Such telephonic survey was done on people and frontline health workers of rural areas to know about vaccination drive and related hesitancy among rural masses.

We divided our survey in to two sections. First section consists of population demographic information including age, sex, occupation etc. We compiled our 11 questions dealing with various related issues of vaccine hesitancy (Supplementary text). We provided multiple options to the respondents towards an objective direction. In second section, participants of telephonic interviews were the frontline health workers including CHO(Community Health Workers), ANM(Auxiliary Nurse Midwives), ASHA (Accredited Social Health Activists) and SANGINI, who had maintained their record and have shared with us their personal observations. The interview was a structured interview and the main emphasis was over the two questions that were asked-

Q.1:- What is the main restraint among people of rural India to participate in COVID-19 vaccination?

Q.2:- How do you see vaccination drive in your area and if you have to suggest few reasons, kindly provide few of them to make vaccination drive more inclusive and widespread? We tend to use it as an additional data to have a better and broadened look over the conclusion been drawn from our study and whether it complies with it or not.

Apart from the health-workers, we also did a second telephonic interview with people from the rural areas. This interview was also structured and it consisted of two questions-

Q.1:- Do you want to get vaccinated?

Q.2:- If not, then why?

## Results and discussion

Our questionnaire was designed in such a way that it should reflect the mass feeling about the nature of virus, the second wave, comments on measure taken by the government during second wave and rumors leads to vaccine hesitancy (Supplementary text). Apart from the highly infective virus variants during the second wave (*16*), the role of public was also concerning. A large proportion of participants (0.75) either relied on rumors or did not the take the virus seriously. And when asked that what sort of steps, according to the respondents, would help in the prevention of the coronavirus, majority of the people think that total vaccination and personal consciousness (0.658) will be a better tool than the lockdowns (Supplementary text).

An interesting result which our study yielded is that in some questions, there was a significant difference (two tailed p value <0.001) (in the responses between male and female respondents (Supplementary Fig. 1). For example, significantly lower number of females think that the coronavirus is lab made or biological weapon, whereas more number of females think COVID-19 as natural pandemic (two tailed p value <0.001) (Supplementary text). Similarly, more number of females followed the health instructions than the males (two tailed p value <0.001).

During the survey, we have found that many of the participants had an impression that vaccine has some relation with the introduction of second wave. Therefore, first we have investigated the vaccine hesitation during the second wave (Supplementary Fig. 1). We have looked to the data of vaccination during March-June 2021 (*11*). We found a major fall of the vaccination during the second wave. The most important reason of this fall was vaccine hesitancy due to the fear of the second wave. While talking with the people during the survey, we have found that as soon as second wave started to spread people run for the vaccine. This has created a large rush to the vaccination centers. Many of them have been infected due to large gathering at the vaccination centers. This has created a confusion in the society that the people are being infected after taking vaccines (Supplementary Fig. 1 and Supplementary text). Thus, vaccines are not helping to stop the infection. Spread of this rumor through word-of-mouth has reduced the vaccinations significantly (two tailed p value <0.0001) (*11*). During the second wave when the test positive rate was on the peak, the daily vaccination was low. However, it has to be understood that it takes 3-4 weeks to develop the antibodies after the vaccination (*17*).

From our research, we found that though a large proportion of people (0.849) would prefer to get vaccinated, are aware of vaccines (0.962) and know that the vaccine for SARS-CoV-2 prevents COVID-19 (0.899), a substantial proportion of people reported that they would refrain to get vaccinated (0.15; Fig. 1). Interestingly, the proportion of vaccine hesitancy was found similar for both male and female participants.

**Figure 1.**
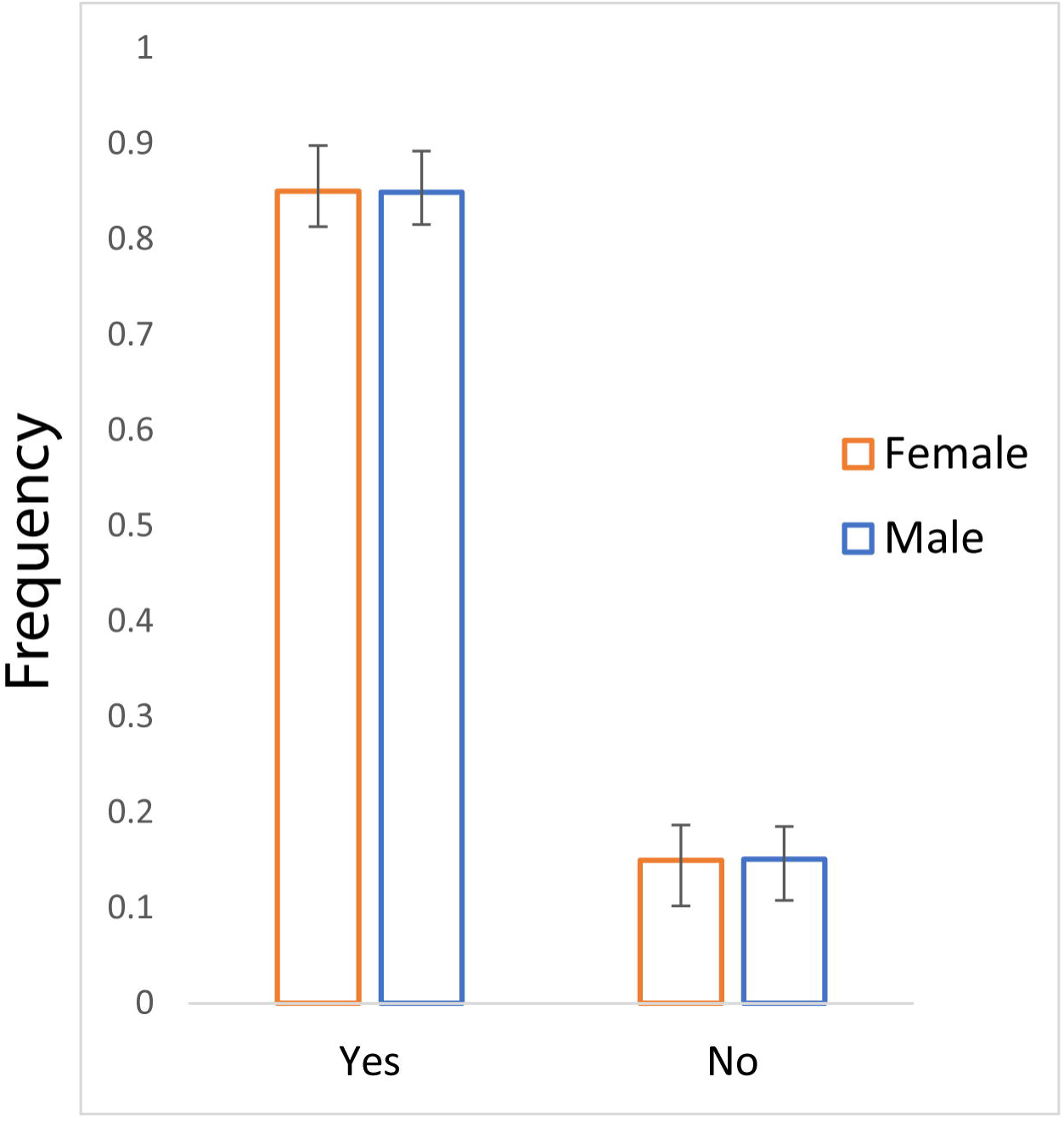
The bar plot of frequency (with 95%CI) showing vaccine hesitancy for male and female participants.

So far, in the SARS-CoV-2 evolution, we have seen that this virus can create more hazardous variants with the time (*16*). And we are fortunate that no variant has been found that completely evades the vaccine induced immunity, but with large number of vaccination, a non-vaccinated pool may provide a reservoir for the virus to multiply and mutate. Thus, it may provide an opportunity to emerge new variants. Moreover, the selection pressure on virus against the background of a largely vaccinated population, may favor a variant that will be resistant to the vaccine. Therefore, the real danger is from those people who have never been vaccinated or infected before, such people will provide ground for a new variant of the virus and if it develops immunity to the vaccine, then it will be impossible to control the epidemic. The progress we have made against this pandemic will be lost.

In our knowledge, our study is perhaps the first attempt to systematically bring out some potential factors related to the COVID-19 vaccine hesitancy among North Indians. However, a retrospective study following face-to-face or telephonic structured interview with qualitative approach such as thematic analysis may bring further insight into the dynamics of vaccine hesitancy among Indians. Similarly, the post second wave status of vaccine hesitancy also need to be explored using the same interview format and contrasted with the retrospective data to understand the extent of vaccine hesitancy and changing factors. Since we have used a structured questionnaire with predefined response format, the study is fraught with danger of subjective biases of the researchers as the proposed factors for vaccine hesitancy were limited by the researchers. The open ended questions for listing the reasons for vaccine hesitancy may bring newer insights as well as additional factors of vaccine hesitancy that could not be foreseen by us while framing the response to the question of vaccine hesitancy.

## Data Availability

All data produced in the present work are contained in the manuscript.

## Acknowledgements

This work is supported by the Faculty IOE grant BHU (6031). GC is supported by SERB India (CRG/2018/001727), PPS is supported by CSIR fellowship, Govt. of India.

## Author contributions

GC, US, AT, JK, SD, SV, VS, DD, PPS and VR conceived and designed this study. US, AT, JK, SD, SV, VS, DD, PPS collected the data and also conducted the online and offline surveys. PPS, GC, DD, PK and VR analyzed the data. GC, US, AT, JK, SD, SV, DD, and RP wrote the manuscript from the inputs of other co-authors. All authors contributed to the article and approved the submitted version.

## Data Availability Statement

All datasets generated for this study are included in the article/Supplementary Material.

## Competing interests

The authors declare no competing interests.

## Conflict of Interest

Authors declare that the research was conducted in the absence of any commercial or financial relationships that could be construed as a potential conflict of interest.

## Notes

### Competing Interest Statement

The authors have declared no competing interest.

### Author Declarations

The study has been approved by the Ethical committees of Banaras Hindu University, Varanasi and VBS Purvanchal University, Jaunpur, India.

## References

1. O. Batiha, T. Al-Deeb, E. Al-zoubi, E. Alsharu, Impact of COVID-19 and other viruses on reproductive health. Andrologia. 52, e13791 (2020).

2. B. B. Finlay, K. R. Amato, M. Azad, M. J. Blaser, T. C. Bosch, H. Chu, M. G. Dominguez-Bello, S. D. Ehrlich, E. Elinav, N. Geva-Zatorsky, The hygiene hypothesis, the COVID pandemic, and consequences for the human microbiome. Proc. Natl. Acad. Sci. 118 (2021).

3. J. BrainardMay. 13, 2020, 12:15 Pm, Scientists are drowning in COVID-19 papers. Can new tools keep them afloat? Sci. AAAS (2020), (available at https://www.sciencemag.org/news/2020/05/scientists-are-drowning-covid-19-papers-can-new-tools-keep-them-afloat).

4. J. A. Singh, R. Ravinetto, COVID-19 therapeutics: how to sow confusion and break public trust during international public health emergencies. J. Pharm. Policy Pract. 13, 1–7 (2020).

5. S. Crawford, Gender-related Irritability, Confusion, Anger, and Frustration Associated with COVID-19 Infection and Mortality. J. Res. Gend. Stud. 10, 138–147 (2020).

6. India coronavirus dispatch: AYUSH recommendations add to confusion on norms | Business Standard News, (available at https://www.business-standard.com/article/current-affairs/india-coronavirus-dispatch-ayush-recommendations-add-to-confusion-on-norms-120110501015_1.html).

7. A. Agrawal, S. Kolhapure, A. Di Pasquale, J. Rai, A. Mathur, Vaccine Hesitancy as a Challenge or Vaccine Confidence as an Opportunity for Childhood Immunisation in India. Infect. Dis. Ther. 9, 421–432 (2020).

8. J. Jain, S. Saurabh, P. Kumar, M. K. Verma, A. D. Goel, M. K. Gupta, P. Bhardwaj, P. R. Raghav, COVID-19 vaccine hesitancy among medical students in India. Epidemiol. Infect., 1–28 (2021).

9. J. C. Lyu, E. Le Han, G. K. Luli, COVID-19 vaccine–related discussion on Twitter: topic modeling and sentiment analysis. J. Med. Internet Res. 23, e24435 (2021).

10. S. Praveen, R. Ittamalla, G. Deepak, Analyzing Indian general public’s perspective on anxiety, stress and trauma during Covid-19-a machine learning study of 840,000 tweets. Diabetes Metab. Syndr. Clin. Res. Rev. 15, 667–671 (2021).

11. Coronavirus in India: Latest Map and Case Count, (available at https://www.covid19india.org).

12. M. S. N. D. March 23, 2021UPDATED: March 23, 2021 17:25 Ist, India to vaccinate all above 45 from April 1. India Today, (available at https://www.indiatoday.in/coronavirus-outbreak/vaccine-updates/story/india-to-vaccinate-people-above-45-from-april-1-1782661-2021-03-23).

13. R. Ranjan, A. Sharma, M. K. Verma, Characterization of the Second Wave of COVID-19 in India. medRxiv (2021).

14. P. Asrani, M. S. Eapen, M. I. Hassan, S. S. Sohal, Implications of the second wave of COVID-19 in India. Lancet Respir. Med. (2021).

15. Misinformation by Oppn led to vaccine hesitancy in India: Union minister Puri. Hindustan Times (2021), (available at https://www.hindustantimes.com/india-news/misinformation-by-oppn-led-to-vaccine-hesitancy-in-india-union-minister-puri-101631895943676.html).

16. K. Kupferschmidt, M. Wadman, Delta variant triggers new phase in the pandemic (2021).

17. P. Gupta, P. P. Singh, A. Pathak, V. N. Mishra, RE: Prior SARS-CoV-2 infection produce adequate antibodies after first vaccine dose (2021).

