## Supplementary text for "Vaccine hesitancy for coronavirus SARS-CoV-2 in North India"

**Q.1 - What is coronavirus?**

| **Options** | **Total (n=727)** | **Male (n=425)** | **Female (n=302)** |
| --- | --- | --- | --- |
| Natural pandemic | 0.693 (0.726-0.659) | 0.631(0.584-0.675) | 0.781 (0.731-0.824) |
| Lab made virus | 0.180 (0.154-0.21) | 0.224(0.187-0.266) | 0.119 (0.087-0.161) |
| Biological weapon | 0.073 (0.056-0.094) | 0.096(0.072-0.128) | 0.040 (0.023-0.068) |
| Global conspiracy | 0.034 (0.023-0.050) | 0.0310.018-0.052) | 0.040 (0.023-0.068) |
| Government weapon | 0.019 (0.012-0.032) | 0.019(0.01-.0037) | 0.020 (0.009-0.043) |

**Q.2 -What does corona vaccine do?**

| **Options** | **Total (n=533)** | **Male (317)** | **Female (216)** |
| --- | --- | --- | --- |
| Makes you impotent | 0.006 (0.002-0.016) | 0.003 (0.001-0.017) | 0.009 (0.003-0.033) |
| Prevents corona | 0.899 (0.87-0.921) | 0.905 (0.868-0.933) | 0.889 (0.84-0.924) |
| Population control | 0.019 (0.01-0.034) | 0.019 (0.009-0.041) | 0.019 (0.008-0.047) |
| Makes you emotionless | 0.019 (0.01-0.034) | 0.019 (0.009-0.041) | 0.019 (0.008-0.047) |
| Leads to death | 0.058 (0.041-0.081) | 0.054 (0.034-0.084) | 0.065 (0.039-0.106) |

**Q.3 - What was the role of the government during the corona pandemic?**

| **Options** | **Total (n=813)** | **Male (n=482)** | **Female (n=331)** |
| --- | --- | --- | --- |
| Can be improved | 0.389 (0.356-0.423) | 0.351 (0.309-0.394) | 0.444 (0.392-0.498) |
| Irresponsible attitude | 0.219 (0.192-0.249) | 0.241 (0.205-0.281) | 0.187 (0.149-0.233) |
| Satisfactory | 0.097 (0.0179-0.119) | 0.087 (0.065-0.116) | 0.112 (0.082-0.15) |
| Very good | 0.111 (0.091-0.134) | 0.116 (0.091-0.148) | 0.103 (0.075-0.14) |
| Worrying | 0.185 (0.159-0.213) | 0.205 (0.172-0.244) | 0.154 (0.119-0.197) |

**Q.4 - What was the role of the public in spread of coronavirus (SARS-CoV-2)?**

| **Options** | **Total (n=848)** | **Male (n=480)** | **Female (n=368)** |
| --- | --- | --- | --- |
| Relied on rumors | 0.317 (0.287-0.349) | 0.335 (0.295-0.379) | 0.293 (0.249-0.342) |
| Agreed with government | 0.083 (0.066-0.103) | 0.085 (0.064-0.114) | 0.079 (0.056-0.111) |
| Followed health instructions | 0.123 (0.102-0.146) | 0.081 (0.06-0.109) | 0.177 (0.141-0.219) |
| Didn’t take seriously | 0.433 (0.4-0.466) | 0.465 (0.420-0.509) | 0.391 (0.343-0.442) |
| Took seriously | 0.039 (0.028-0.054) | 0.033 (0.021-0.053) | 0.060 (0.040-0.089) |

**Q.5 - Which of the following steps according to you would help to stop the infection of coronavirus?**

| **Options** | **Total (n=1218)** | **Male (n=721)** | **Female (n=497)** |
| --- | --- | --- | --- |
| Total lockdown | 0.250 (0.226-0.275) | 0.247 (0.217-0.280) | 0.256 (0.219-0.296) |
| Partial lockdown | 0.089 (0.075-0.107) | 0.097 (0.078-0.121) | 0.078 (0.058-0.106) |
| Personal consciousness and awareness | 0.380 (0.353-0.407) | 0.368 (0.333-0.403) | 0.400 (0.358-0.444) |
| Total vaccination | 0.278 (0.254-0.304) | 0.288 (0.257-0.323) | 0.266 (0.229-0.306) |

**Q.6 – How do you view the health management of India during COVID-19 second wave?**

| **Options** | **Total (n=656)** | **Male (n=370)** | **Female (n=286)** |
| --- | --- | --- | --- |
| Good | 0.064 (0.048-0.085) | 0.065 (0.044-0.095) | 0.063 (0.040-0.097) |
| Very Good | 0.046 (0.032-0.065) | 0.041 (0.025-0.066) | 0.052 (0.032-.085) |
| Satisfactory | 0.168 (0.141-0.198) | 0.157 (0.123-0.197) | 0.182 (0.141-0.231) |
| Unsatisfactory | 0.410 (0.373-0.448) | 0.422 (0.372-0.473) | 0.395 (0.340-0.453) |
| Average | 0.313 (0.278-0.349) | 0.316 (0.271-0.365) | 0.308 (0.257-0.364) |

**Q.7 - Would you prefer to get vaccinated?**

| **Options** | **Total (n=603)** | **Male (n=337)** | **Female (n=266)** |
| --- | --- | --- | --- |
| Yes | 0.849 (0.818-0.875) | 0.849 (0.806-0.883) | 0.850 (0.802-0.887) |
| No | 0.151 (0.125-0.182) | 0.151 (0.117-0.194) | 0.150 (0.113-0.198) |

**Q.9 - Did you take the Covid test?**

| **Options** | **Total (n=603)** | **Male (n=337)** | **Female (n=266)** |
| --- | --- | --- | --- |
| Yes | 0.388 (0.350-0.428) | 0.418 (0.367-0.472) | 0.350 (0.295-0.409) |
| No | 0.612 (0.572-0.650) | 0.582 (0.528-0.633) | 0.650 (0.591-0.705 ) |

**Q.9a - What was the test result?**

| **Options** | **Total (n=317)** | **Male (n=175)** | **Female (n=142)** |
| --- | --- | --- | --- |
| Positive | 0.129 (0.097-0.179) | 0.183 (0.133-0.247) | 0.063 (0.034-0.116) |
| Negative | 0.871 (0.829-0.903) | 0.817 (0.753-0.867) | 0.937 (0.884-0.966) |

**Q.10 - Which vaccine are you aware of?**

| **Options** | **Total (n=1274)** | **Male (n=759)** | **Female (n=515)** |
| --- | --- | --- | --- |
| All | 0.038 (0.029-0.050) | 0.025 (0.016-0.039) | 0.056 (0.40-0.080) |
| Covishield | 0.349 (0.323-0.375) | 0.358 (0.325-0.393) | 0.334 (0.295-0.376) |
| Covaxin | 0.376 (0.350-0.403) | 0.364 (0.330-0.398) | 0.394 (0.353-0.437) |
| Sputnik-V | 0.238 (0.215-0.262) | 0.253 (0.223-0.285) | 0.216 (0.182-0.253) |

**Q.11 - Will vaccination prevent COVID-19 lifelong?**

| **Options** | **Total (n=632)** | **Male (n=278)** | **Female (n=351)** |
| --- | --- | --- | --- |
| Yes | 0.082 (0.063-0.106) | 0.119 (0.086-0.162) | 0.054 (0.035-0.083) |
| No | 0.441 (0.403-0.480) | 0.572 (0.513-0.629) | 0.342 (0.294-0.393) |
| Not Sure | 0.472 (0.433-0.511) | 0.572 (0.513-0.629) | 0.396 (0.346-0.448) |
