## Supplementary figures and images for "Vaccine hesitancy for coronavirus SARS-CoV-2 in North India"

### Supplementary Figure 1

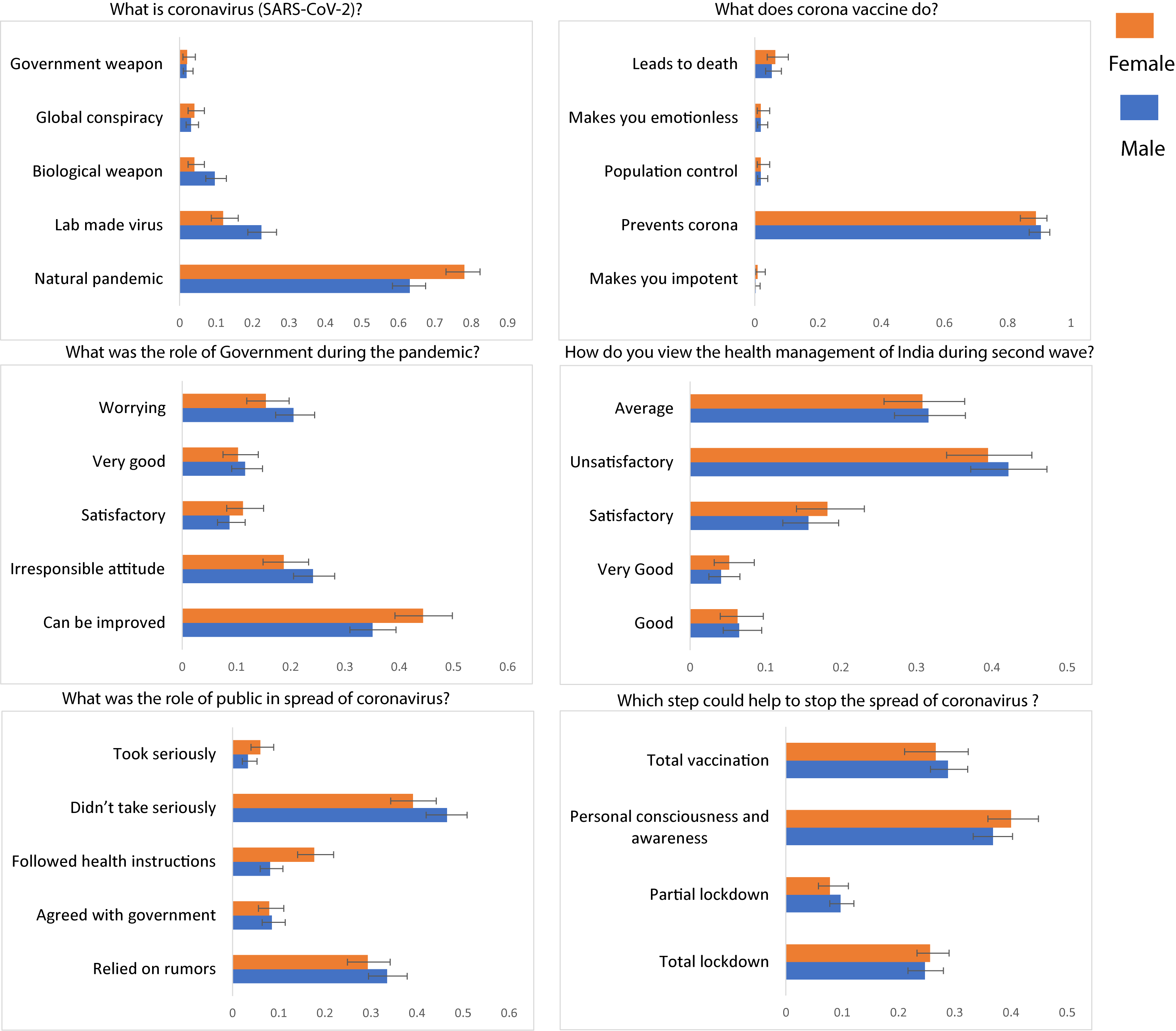
